# A validation of machine learning-based risk scores in the prehospital setting

**DOI:** 10.1101/19007021

**Authors:** Douglas Spangler, Thomas Hermansson, David Smekal, Hans Blomberg

## Abstract

**Background:** The triage of patients in pre-hospital care is a difficult task, and improved risk assessment tools are needed both at the dispatch center and on the ambulance to differentiate between low- and high-risk patients. This study develops and validates a machine learning-based approach to predicting hospital outcomes based on routinely collected prehospital data.

**Methods:** Dispatch, ambulance, and hospital data were collected in one Swedish region from 2016 - 2017. Dispatch center and ambulance records were used to develop gradient boosting models predicting hospital admission, critical care (defined as admission to an intensive care unit or in-hospital mortality), and two-day mortality. Model predictions were used to generate composite risk scores which were compared to National Early Warning System (NEWS) scores and actual dispatched priorities in a similar but prospectively gathered dataset from 2018.

**Results:** A total of 38203 patients were included from 2016-2018. Concordance indexes (or area under the receiver operating characteristics curve) for dispatched priorities ranged from 0.51 – 0.66, while those for NEWS scores ranged from 0.66 - 0.85. Concordance ranged from 0.71 – 0.80 for risk scores based only on dispatch data, and 0.79 – 0.89 for risk scores including ambulance data. Dispatch data-based risk scores consistently outperformed dispatched priorities in predicting hospital outcomes, while models including ambulance data also consistently outperformed NEWS scores. Model performance in the prospective test dataset was similar to that found using cross-validation, and calibration was comparable to that of NEWS scores.

**Conclusions:** Machine learning-based risk scores outperformed a widely-used rule-based triage algorithm and human prioritization decisions in predicting hospital outcomes. Performance was robust in a prospectively gathered dataset, and scores demonstrated adequate calibration. Future research should investigate the generality of these results to prehospital triage in other settings, and establish the impact of triage tools based on these methods by means of randomized trial.

## Introduction

Emergency care systems in the developed world face increasing burdens due to an aging population [1–4], and in prehospital care it is often necessary to prioritize high-risk patients in situations where resources are scarce. Prehospital care systems have also increasingly sought to identify patients not in need of emergency care, and to direct these patients to appropriate forms of alternative care both upon contact via telephone with the dispatch center, and upon the arrival of an ambulance to a patient [5–12]. Performing these tasks safely and efficiently requires not only well-trained prehospital care providers and carefully considered clinical guidelines, but also the employment of triage algorithms able to perform risk differentiation across the diverse cohort of patients presenting to prehospital care systems.

Systems to differentiate high- and low-risk patients in prehospital care have typically relied on rule-based algorithms. Many common algorithms seek to identify specific high-acuity conditions within certain subsets of patients such as cardiac arrest, trauma, or stroke [13–15]. Other algorithms are intended for use within a broader cohort of patients, including Critical Illness Prediction (CIP) scores and the National Early Warning System (NEWS) [16–19], and the Medical Priority Dispatching System (MPDS) [20] for Emergency Medical Dispatching (EMD). In applying such tools, providers commonly “over-triage” patients, as false negatives are thought to be associated with far greater costs than false positive findings [21–24]. In the context of trauma care, the American College of Surgeons Committee on Trauma (ACS-CoT) recommend that decision rules to identify patients suitable for direct transport to a level-1 trauma center have a sensitivity of 95%, while an appropriate level of specificity may be as low as 65% [24,25]. We identified no guidelines establishing appropriate levels of sensitivity for decision rules intended to identify patients suitable for referral to alternate forms of care by prehospital care providers. Given the costs of missing true emergencies in this application, the required level of sensitivity may be similarly high.

In the context of Emergency Department (ED) triage, Machine Learning (ML) based triage algorithms can out-perform their rule-based counterparts in predicting general measures of patient outcome [26–29]. We identified no research relating to the ability of prehospital data to similarly predict hospital outcomes, though there are indications that ML techniques may be effective in identifying specific high-acuity conditions such as cardiac arrest at the dispatch center [30]. ML-based approaches offer the potential to integrate large and complex sets of predictors, and automatically calculate risk scores for use by care providers. By using prehospital data to predict hospital outcomes, it may be possible to enhance the ability of prehospital care providers to safely identify patients not in need of hospital care. Such low-risk patients could then be directed to less intensive forms of care (e.g. transport to a primary care facility or a home visit by a mobile care physician), thus alleviating the increasingly vexing problem of overcrowding at EDs [31–33]. Such scores could also be used to improve the overall accuracy of ambulance dispatching systems, ensuring that high-risk patients are prioritized over those with less need for emergency care.

In this study, we developed machine learning models to predict patient outcomes in a broad cohort of patients at two distinct points in the chain of emergency care: In the EMD center prior to ambulance dispatch, and on the ambulance after making contact with the patient. We investigated the feasibility of using these methods to improve the decisional capacity prehospital care providers in these settings by comparing their accuracy with a previously validated triage algorithm (NEWS), and with prioritization decisions made by nurses at the EMD center per current clinical practice.

## Methods

### Source of Data

This study took place in the region of Uppsala, Sweden, with a size of 8 209 km2, and a population of 376 354 in 2018. The region is served by two hospital-based EDs, a single regional EMD center staffed by Registered Nurses (RNs) employing a self-developed Clinical Decision Support System (CDSS), and 18 RN-staffed ambulances. The CDSS consists of an interface wherein dispatchers first seek to identify a set life-threatening conditions (cardiac/respiratory arrest or unconsciousness), and then document the primary complaint of the patient. Based on the documented complaint, a battery of questions is presented, the answers to which determine the priority of the call, or open additional complaints. While the specific set of questions are idiosyncratic to this and 3 other Swedish regions, its structure is similar to other dispatch CDSS such as the widely-used MPDS [20].

Ambulance responses are triaged by an RN to one of four priority levels, with 1A representing the highest priority calls (e.g. cardiac/respiratory arrest), and 1B representing less emergent calls still receiving a “lights and sirens” (L&S) response. Calls with a priority of 2A represent urgent, but non-emergent ambulance responses, while 2B calls may be held to ensure resource availability.

Records from January 2016 to December 2017 were extracted to serve as the basis for all model development. Upon finalizing the methods to be reported upon, records from January to December 2018 were extracted to form a test dataset to investigate the prospective performance of the models. The data in this study were extracted from databases owned by the Uppsala ambulance service containing dispatch, ambulance, and hospital outcome data collected routinely for quality assurance and improvement purposes. Ambulance records were deterministically linked to dispatch records based on unique record identifiers available in both systems. Hospital records were extracted from the regional Electronic Medical Records (EMR) system based on patient Personal Identification Numbers (PINs) collected either by dispatchers or ambulance crews. This study was approved by the Uppsala regional ethics review board (dnr 2018/133).

### Participants

Inclusion and exclusion criteria were defined so as to enable comparison with other studies of ML based triage systems in the ED, and with previously validated risk assessment instruments. All dispatch records associated with a primary ambulance response to a single-patient incident (i.e., excluding multi-patient traffic accidents and planned inter-facility transports) were selected for inclusion. Records lacking documentation in the CDSS used at the EMD center were excluded, as were records in which an invalid PIN or multiple PINs were documented. Dispatch records with no associated ambulance journal (e.g. calls cancelled *en route*, or where no patient was found), and records indicating that the patient was treated and left at the scene of the incident were excluded. We further excluded records where no EMR system entry associated with the patient at the appropriate time could be identified (typically due to documentation errors, or transports to facilities outside of the studied region), and EMR system records indicating that the patient was transported to a non-ED destination (e.g. a primary/urgent care facility, or a direct admission to a hospital ward). We also excluded patients with ambulance records missing measurements of more than two of the vital signs necessary to calculate a NEWS score. Patients under the age of 18 were excluded as NEWS scores are not valid predictors of risk for pediatric patients.

### Outcomes

We selected three outcome measures based on their face validity in representing a range of outcome acuity levels, and based to their use in previous studies; 1) patient admission to a hospital ward [26–28,34], 2) the provision of critical care, defined as admission to an Intensive Care Unit (ICU) or in-hospital mortality [26,28], and 3) all-cause patient mortality within two days [18,19].

While each of these outcomes represent an important aspect of the overall risks associated with a patient, no single outcome measure was thought to provide a full picture of patient acuity. As such, we chose to combine these outcomes by predicting the likelihood of each outcome occurring independently, and then combining predictions into a single composite risk score. This is a novel approach, as previous researchers have either investigated only single measures of patient outcome [27,28], or binned scores across specific ranges of predicted likelihoods [26,34]. The method we propose results in composite risk scores reflecting the normalized mean likelihood of several outcomes with face validity as being representative of patient acuity occurring, without incurring the loss of information associated with binning continuous variables. We applied no weights in the compositing process, as the relative importance of these measures in in establishing the overall acuity of the patient is not known.

### Predictors

Predictors extracted from the dispatch system included patient demographics (age and gender), the operational characteristics of the call (Hour and month that the call was received, haversine distance to the nearest ED, and prior contacts with the EMD center by the patient), and the clinical characteristics of the call as documented in the existing rule-based CDSS. We included the 59 complaint categories, and the 1592 distinct question and answer combinations available in the CDSS as potential predictors in our models. Each of the questions in the CDSS was encoded with a 1 representing a positive answer to the question, and 0 representing a negative answer to the question. Questions with multiple potential answers were encoded on a numerical scale in cases where the answers were ordinal (e.g., “How long have the symptoms lasted?”), and as dummy variables if the answers were non-ordered. The recommended priority of the call based on the existing rule-based triage system was also included as a predictor in the dispatch dataset.

Predictors extracted from ambulance records represented the information which would be available at the time of patient hand-over to ED staff, and included the primary and secondary complaints, additional operational characteristics (times to reach the incident, on scene, and to the hospital), vital signs, patient history, medications and procedures administered, and the clinical findings of ambulance staff. Descriptive statistics for the included predictors are reported in S1 Table.

To provide a basis for comparison, we extracted the dispatched priority of the call as determined by the RN handling the call at the EMD center, and retrospectively calculated NEWS scores for each included patient. If multiple vital sign measurements were taken, the first set was used both as model predictors and to calculate NEWS scores.

### Missing data

Missing vital sign measurements in ambulance records are not likely to be missing completely at random, and must be considered carefully [35,36]. Based on exploratory analysis and clinical judgement, we surmised that records missing at most two of the vital signs constituting the NEWS score fulfilled the missing at random assumption necessary to perform multiple imputation. Missing vitals were multiply imputed five times using predictive mean matching over 20 iterations as implemented in the ‘mice’ R package [37]. The characteristics of the imputed data were examined, and we chose to use the median of the imputed vital signs to calculate NEWS scores. Multiply imputed data were not used as predictors, with missing data handled natively by the ML models used here.

### Statistical analysis

We entered each set of predictors transformed as previously described into gradient boosting models as implemented in the XGBoost R package [38]. This algorithm involves the sequential estimation of multiple weak decision trees, with each additional tree reducing the error associated with the previously estimated trees [39]. Model predictions were combined into composite risk scores by scaling each set of outcome predictions to have a population mean of zero and a standard deviation of one. These were then averaged and a log transformation was applied to improve calibration, resulting in a composite risk score following a normal distribution.

We investigated model discrimination using Receiver Operating Characteristics (ROC) curves, using the area under these curves (a measure equivalent to the concordance index, or c-index of the model) as summary performance measures [39].

Precision/Recall curves and their corresponding areas under the curve are included in S2 Analysis. 95% confidence intervals for descriptive statistics and c-index values were generated based on the percentiles of 1000 basic bootstrap samples (using stratified resampling for c-index values) as implemented in the ‘boot’ R package [40]. Model calibration overall and in a number of sub-populations was investigated visually using lowess smoothed calibration curves, and summarized using the mean absolute error between predicted and ideally calibrated probabilities using the ‘val.prob’ function from the ‘rms’ R package [41].

We considered the performance of the models in the prospective dataset to be the best metric of future model performance, though results in this field have previously been reported based on cross-validation [26,34] or randomly selected hold-out samples [28]. In this paper we report our main findings based on model performance in a prospective test dataset, an include results based on cross-validation for comparison. Model performance in the training dataset was estimated using 5-fold cross-validation (CV), and model performance in the testing dataset was based on models estimated using the full training dataset.

Readers interested in further details of the methods employed to produce the results reported here are encouraged to peruse the commented source code found in S6 Code. All model development and validation was performed using R version 3.5.3 [42].

## Results

### Participants

A total of 68 668 records were collected, of which 45 045 were in the training dataset, and 23 623 were in the test dataset as reported in Table 1. Overall, 30 465 records (44%) were excluded due all criteria. A lower proportion of records were excluded from the test dataset, primarily due to fewer non-matched ambulance and hospital records.

**Table 1.** Results of applying exclusion criteria.

|  | Training dataset (2016-2017) |  |  | Test dataset (2018) |  |  |
| --- | --- | --- | --- | --- | --- | --- |
|  | Excluded,<br>N | Excluded,<br>percent | Remaining,<br>N | Excluded,<br>N | Excluded,<br>percent | Remaining,<br>N |
| Original |  |  | 45045 |  |  | 23623 |
| No dispatch CDSS data | 2358 | 5.5 | 42687 | 857 | 3.8 | 22766 |
| Missing PIN | 2113 | 5.2 | 40574 | 1244 | 5.8 | 21522 |
| No ambulance journal | 2526 | 6.6 | 38048 | 933 | 4.5 | 20589 |
| No ambulance transport | 3879 | 11.4 | 34169 | 2461 | 13.6 | 18128 |
| No hospital journal | 3958 | 13.1 | 30211 | 1429 | 8.6 | 16699 |
| No ED visit | 2939 | 10.8 | 27272 | 1590 | 10.5 | 15109 |
| Missing > 2 vitals | 1336 | 5.2 | 25936 | 829 | 5.8 | 14280 |
| Patient age < 18 | 1328 | 5.4 | 24608 | 685 | 5 | 13595 |
| Final | 20437 | 45.4 | 24608 | 10028 | 42.5 | 13595 |

Summary statistics describing the characteristics of all patients included in the study (across both training and testing sets), both in total and stratified by dispatched priority are presented in table 2. We found that ambulance predictors and outcomes were generally distributed such that higher priority calls had higher levels of patient acuity, with the notable exception of hospital admission which remained constant at around 50% regardless of dispatched priority. Higher priority patients were generally younger, more often male, and had a higher proportion of missing vital signs. Overall, at least one vital sign was missing in a quarter of ambulance records, with the most commonly missing vital sign measurement being the patient’s body temperature. Temperature was missing in 15% of cases, and other vital signs were missing in less than 5% of cases as reported in S1 Table. Multiple imputation of these vital signs resulted in good convergence and similarity to non-imputed data, and NEWS scores based on sets of imputed scores did not differ significantly in terms of predictive value.

**Table 2.** Descriptive statistics of included population.

|  | Priority |  |  |  |  |
| --- | --- | --- | --- | --- | --- |
|  | 1A | 1B | 2A | 2B | Total |
| N | 1283 | 15533 | 17227 | 4160 | 38203 |
| Age, mean | 56.2<br>(54.8-57.4) | 64.5<br>(64.1-64.8) | 67.5<br>(67.2-67.8) | 67.3<br>(66.6-67.9) | 65.9<br>(65.7-66.1) |
| Female, percent | 46.1<br>(43.3-48.9) | 49.4<br>(48.6-50.1) | 53.9<br>(53.2-54.6) | 54.5<br>(52.9-56.0) | 51.9<br>(51.4-52.4) |
| Transported L&S, percent | 38.7<br>(36.0-41.5) | 24.6<br>(23.9-25.2) | 4.3<br>(4.0-4.6) | 2.2<br>(1.7-2.6) | 13.5<br>(13.1-13.8) |
| Ambulance intervention*, percent | 87.9<br>(86.0-89.6) | 87.4<br>(86.9-87.9) | 71.1<br>(70.5-71.8) | 62.1<br>(60.6-63.5) | 77.3<br>(76.9-77.7) |
| Missing vitals, percent | 33.8<br>(31.4-36.3) | 25.7<br>(25.0-26.4) | 24.4<br>(23.7-25.1) | 23.8<br>(22.5-25.0) | 25.2<br>(24.7-25.6) |
| NEWS value, mean | 5.80<br>(5.60-6.01) | 3.76<br>(3.71-3.83) | 2.97<br>(2.93-3.02) | 2.40<br>(2.32-2.48) | 3.33<br>(3.29-3.36) |
| Prior contacts within 30 days, mean | 0.21<br>(0.17-0.24) | 0.17<br>(0.16-0.18) | 0.17<br>(0.16-0.18) | 0.23<br>(0.21-0.25) | 0.18<br>(0.17-0.18) |
| Intensive Care Unit, percent | 10.0<br>(8.3-11.6) | 3.5<br>(3.2-3.7) | 1.7<br>(1.5-1.8) | 1.6<br>(1.2-2.0) | 2.7<br>(2.5-2.8) |
| In-hospital death, percent | 8.7<br>(7.2-10.3) | 4.0<br>(3.7-4.4) | 3.7<br>(3.4-4.0) | 3.9<br>(3.4-4.5) | 4.0<br>(3.8-4.2) |
| Critical care, percent | 16.1<br>(14.1-18.2) | 6.8<br>(6.4-7.2) | 4.9<br>(4.6-5.2) | 4.9<br>(4.3-5.6) | 6.0<br>(5.8-6.3) |
| Admitted, percent | 51.9<br>(49.2-54.6) | 52.3<br>(51.5-53.1) | 52.3<br>(51.6-53.1) | 49.2<br>(47.7-50.8) | 52.0<br>(51.5-52.4) |
| 2-day mortality, percent | 4.8<br>(3.7-5.9) | 1.6<br>(1.4-1.8) | 0.7<br>(0.6-0.9) | 0.7<br>(0.5-1.0) | 1.2<br>(1.1-1.3) |

*Statistics are reported with their bootstrapped 95% confidence interval*
*\* Interventions include Medication administration, Oxygen administration, IV placement,* *Spinal/longbone immobilization, 12-lead EKG capture/transmission to hospital,* *Transport using lights and sirens (L&S), Hospital pre-arrival notification, and* *administration of CPR.*

### Model performance

Receiver operating characteristics curves across the three hospital outcomes for each of the risk prediction scores, as well as for the dispatched priority of the call are presented in fig 1. We found that for all investigated outcomes, risk scores based on ambulance data outperformed all other instruments investigated. NEWS scores had a greater overall c-index than dispatch data-based models for critical care and two-day morality, but at threshold values corresponding to high levels of sensitivity, dispatch data-based risk predictions provided similar levels of specificity. In predicting critical care, NEWS scores were unable to achieve a level of sensitivity corresponding to ACS-CoT guidelines, with a decision rule based on a NEWS score of 1 or more yielding a sensitivity (and 95% CI) of 0.92 (0.90 - 0.94) and a specificity of 0.24 (0.24 - 0.25). At the same level of sensitivity, the dispatch and ambulance data-based risk score yielded specificities of 0.27 (0.27 - 0.28) and 0.36 (0.35 - 0.37) respectively. With regards to 2-day mortality, a decision rule based on NEWS score of 2 or above yields a sensitivity of 0.95 (0.92 - 0.98), corresponding to the ACS-CoT recommendation, while providing a specificity of 0.41 (0.40 - 0.42). At equivalent levels of sensitivity, the dispatch and ambulance based risk scores provide specificities of 0.28 (0.27 - 0.28) and 0.52 (0.51 - 0.53) respectively.

**Fig 1.**
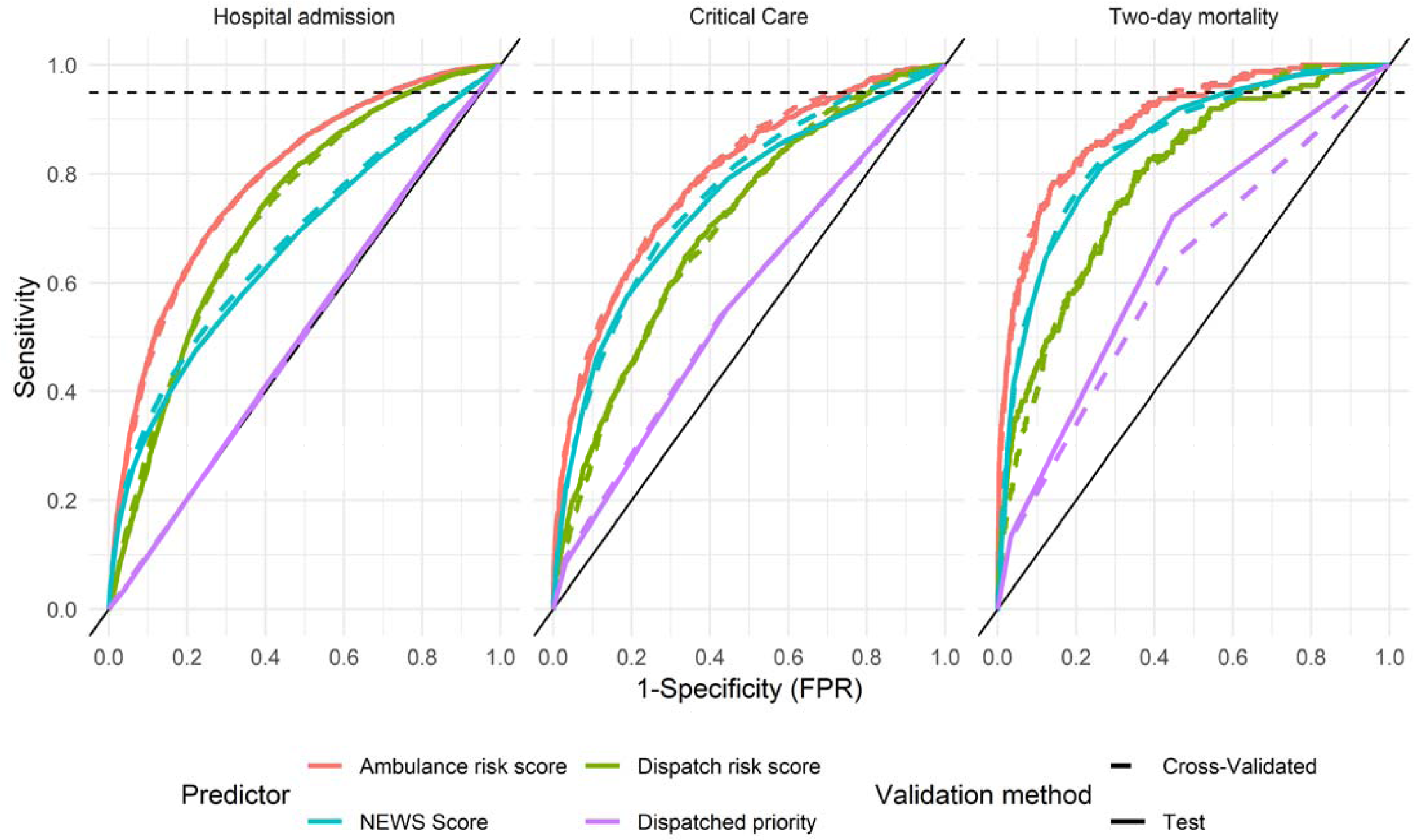
Receiver Operating Characteristics in predicting hospital outcomes. Dotted line corresponds to 95% sensitivity as recommended by the ACS-CoT

Table 3 summarizes the discrimination of the risk assessment instruments for each outcome in the test dataset using the c-index of the model and its 95% confidence interval. ML models based on ambulance data outperformed NEWS scores in terms of c-index for all outcomes. The dispatch data based risk predictions outperformed NEWS in predicting hospital admission, while NEWS scores outperformed the dispatch data based predictions for critical care and two-day mortality in terms of overall discrimination. All risk assessment instruments outperformed dispatched priorities in predicting hospital outcomes, which were found to have some predictive power for critical care and two day mortality, but none for hospital admission. We found no significant differences between model performance using cross-validation and validation in the test dataset.

**Table 3.**
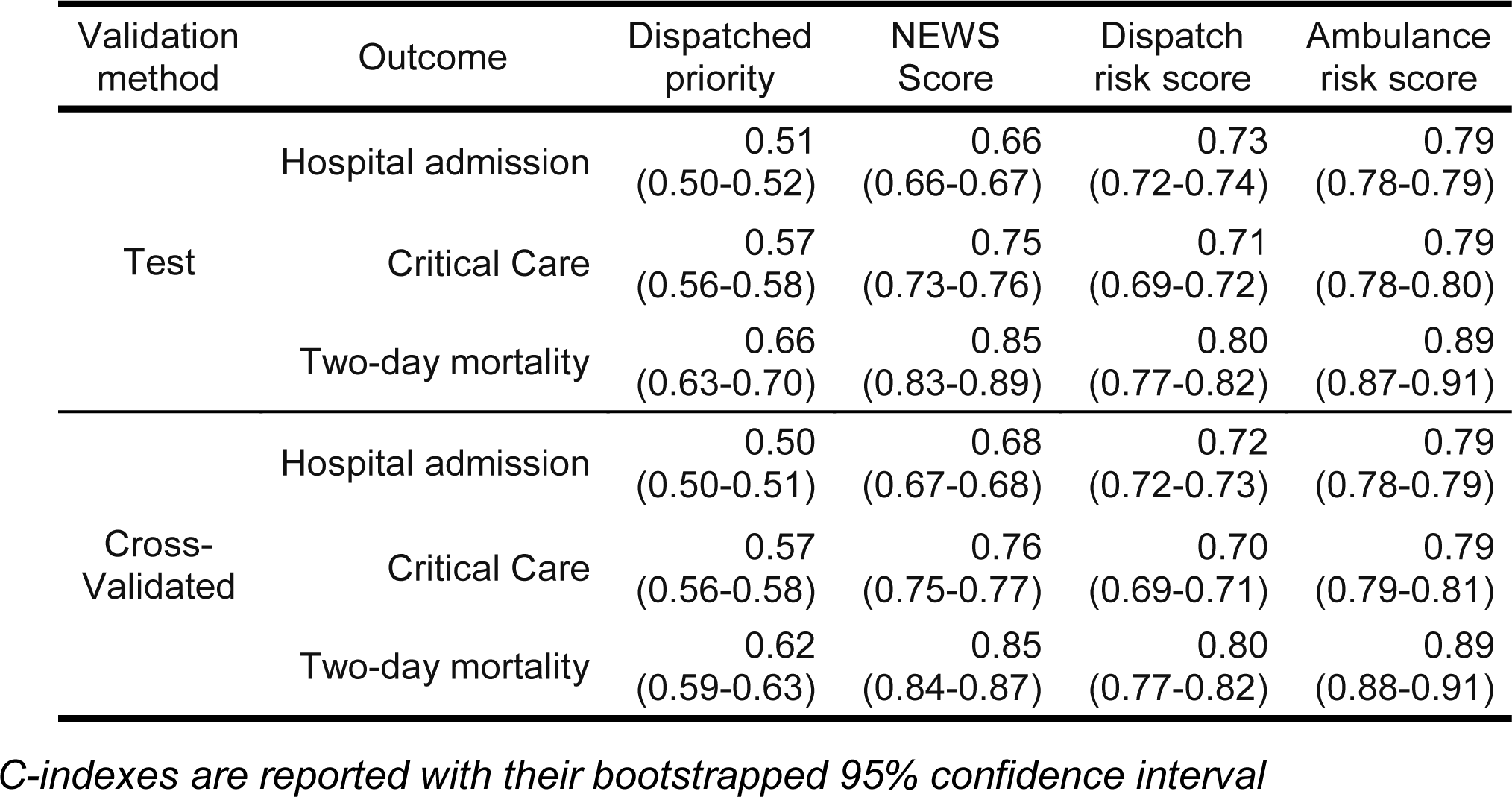
Concordance indexes in predicting hospital outcomes.

| Validation method | Outcome | Dispatched priority | NEWS Score | Dispatch risk score | Ambulance risk score |
| --- | --- | --- | --- | --- | --- |
| Test | Hospital admission | 0.51<br>(0.50-0.52) | 0.66<br>(0.66-0.67) | 0.73<br>(0.72-0.74) | 0.79<br>(0.78-0.79) |
|  | Critical Care | 0.57<br>(0.56-0.58) | 0.75<br>(0.73-0.76) | 0.71<br>(0.69-0.72) | 0.79<br>(0.78-0.80) |
|  | Two-day mortality | 0.66<br>(0.63-0.70) | 0.85<br>(0.83-0.89) | 0.80<br>(0.77-0.82) | 0.89<br>(0.87-0.91) |
| Cross-Validated | Hospital admission | 0.50<br>(0.50-0.51) | 0.68<br>(0.67-0.68) | 0.72<br>(0.72-0.73) | 0.79<br>(0.78-0.79) |
|  | Critical Care | 0.57<br>(0.56-0.58) | 0.76<br>(0.75-0.77) | 0.70<br>(0.69-0.71) | 0.79<br>(0.79-0.81) |
|  | Two-day mortality | 0.62<br>(0.59-0.63) | 0.85<br>(0.84-0.87) | 0.80<br>(0.77-0.82) | 0.89<br>(0.88-0.91) |

*C-indexes are reported with their bootstrapped 95% confidence interval*

We found that both NEWS and ML-based risk scores demonstrated some deviation from ideal calibration as reported in S3 Fig. In terms of mean average error, NEWS scores demonstrated better overall calibration in predicting hospital admission and critical care, but not two-day mortality as reported in S4 Table. In investigating model calibration in sub-populations stratified by age, gender, dispatched priority and patient complaint, some sub-populations did deviate from ideal calibration among both NEWS scores and ML risk scores, though deviations were not consistent across outcomes.

The relative gain in predictive value provided by the 15 most important predictors included in the ambulance data-based models is reported in Fig 2, in order of descending mean gain across the 3 outcomes. Patient age and the provision of oxygen (coded as the liter per minute flow) ranked highest, followed by a number of patient vital signs. Whether or not the patient was transported using lights and sirens to the hospital was a strong predictor of outcomes. A number of measures of call duration (time to the hospital, time on-scene, and time between call receipt and ambulance dispatch), the distance to the nearest ED, and time of day of the call also ranked highly. A summary of the gain provided by all included variables is provided in S1 Table.

**Fig 2.**
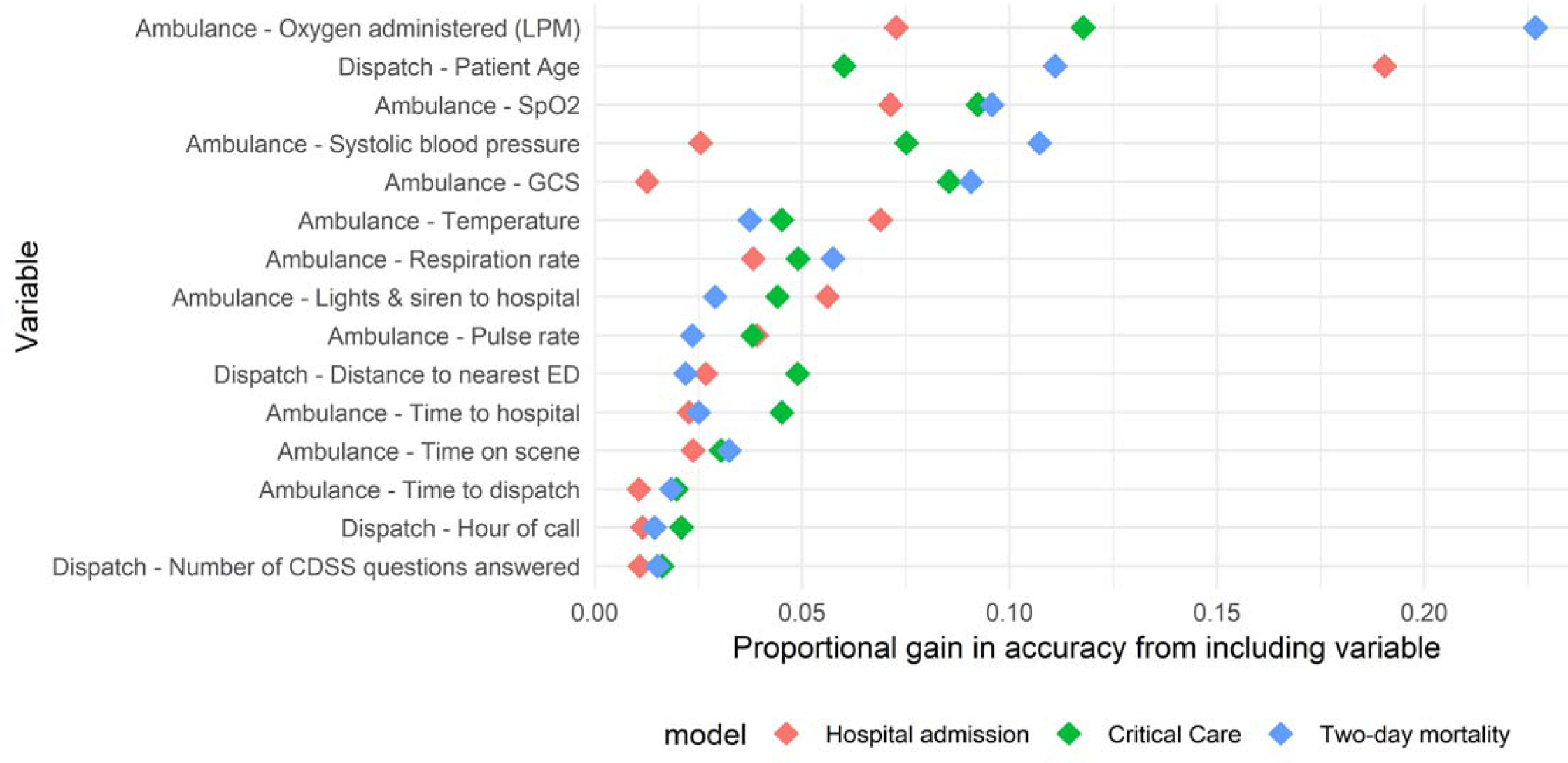
Importance of variables in predicting hospital outcomes in Ambulance models. Variables are arranged in order of descending mean gain across the models predicting the outcomes included in the ambulance data-based risk score

## Discussion

### Limitations

We limited this study to the investigation of a composite score based on an unweighted average of model predictions for three specific hospital outcomes. In doing so, we make the assumption that each of these outcomes is equally important in determining the overall risks associated with the patient. A sensitivity analysis provided in S5 Table demonstrated that while the predictive value of the risk scores did shift in favor of more heavily weighted outcomes across a range of weights, the differences did not impact the main findings of this study. The unweighted average furthermore offered a good compromise in terms of discrimination for each of the constituent outcomes. The most appropriate set of outcomes and associated weights to employ is nevertheless dependent on the intended application of the risk scores, and we recognize that we have examined only one of many potentially valid sets of outcome measures to employ in prehospital risk assessment.

We observed a rate of loss to follow up of around 5-10% upon the application of each of our exclusion criteria. To assess and ameliorate risks associated with data quality issues, we manually spot-checked records to ensure the accuracy of our automated data extraction methods and addressed systematic data extraction issues where we found them, which could account for the lower rate of loss to follow-up we observed in the test dataset as reported in Table 1. The linkage rates found in this study were similar or superior to other studies of prehospital data [43–45]. We also observed c-index values for NEWS scores similar to those found in previous studies; Lane et al. [18] identified c-indexes of 0.85 for NEWS scores in predicting two-day mortality, similar to our value of 0.85 (0.82-0.86). Results were also similar to those identified by Pirneskoski et al. [19], who found a c-index value of 0.84 for NEWS scores in predicting 1-day mortality. Such agreement suggests that the quality of the data in this study is comparable to that of previously published research in the field.

While the ML models reported on in this single-site study performed well in prospective validation, they are not likely to generalize well if applied directly to other contexts. Guidelines regarding hospital admission and intensive care for instance may vary, potentially biasing outcome predictions if these models were applied directly in other settings. Such idiosyncrasies are likely to exist among predictor variables as well: Oxygen was found to have been administered to 17% of patients in this study for instance, a rate which appears to be lower than that found in other contexts [46,47]. In settings where oxygen is administered more liberally, it is not likely to be as strongly associated with patient acuity. The ML framework we employ is however highly flexible, and is likely to produce good results if models were to be trained “from scratch” on other similar datasets. As such, rather than seek to apply the specific models developed in this study to other settings, we encourage researchers to generate and validate novel models based on the framework we propose in other settings. To enhance reproducibility, we sought to adhere to TRIPOD guidelines in reporting our results regarding the development and validation of these models [48], and it is hoped that the source code found in S6 Code will facilitate the replication of-, and improvement upon our results.

### Interpretation

In this study, we found that risk scores generated using ML models based on ambulance data outperformed NEWS scores in predicting hospital outcomes. Risk scores based on data gathered at the EMD center outperformed the prioritizations made by dispatch nurses, and performed comparably to NEWS scores (which are based on physiological data gathered upon patient contact) in settings where high sensitivity is demanded.
Model performance was similar when validated internally using cross-validation and when evaluated in a prospectively gathered dataset, suggesting that the performance of the models is likely to remain stable upon being implemented within the studied context. ML-based risk scores demonstrated acceptable levels of calibration both overall and stratified by age, gender, priority and common call types, and were only mildly sensitive to the selection of alternate sets of weights. Overall, these findings suggest that the application of machine learning methods to routinely collected dispatch and ambulance data is a feasible approach to improving the ability of prehospital care providers to assess the risks associated with their patients in terms of the need for hospital care.

During the development of the methods reported here, we investigated the performance of a number of ML techniques including regularized logistic regression, support vector machines, random forests, gradient boosting, and deep neural networks in the training dataset. As in previous studies [27,49–51], we found that the XGBoost algorithm performed at least as well as other methods we applied to these data in terms of discrimination. We also found that the XGBoost algorithm had several practical benefits, including being invariant to monotonic transformations of the predictors (thus simplifying the data transformation pipeline) [39], and appropriately handling missing data using a sparsity-aware splitting algorithm [38]. While providing good discrimination, the approach does have some drawbacks including being somewhat difficult to interpret, the inability to update models without access to the full original dataset, and that the models are not inherently well calibrated as logistic regression for instance is.

We found the overall calibration of our composite risk scores to be satisfactory, despite their nature as an average of multiple outcomes. Examination of calibration across sub-populations yielded interesting results which could be further examined. We found NEWS scores for instance to systematically under-estimate the probability of hospital admission among older patients - Such miscalibration could be the result of an over-estimation of risks among older patients in the hospital admission process, but could also represent an underlying bias in NEWS scores as currently calculated. Interestingly, all risk scores tended to underestimate the probability of two-day mortality for the oldest quartile of patients. While the usual caution in interpreting post-hoc sub-group analyses is warranted, we found analyses of this type to be useful in developing the models reported here, and in considering how to proceed with their application to clinical practice.

While dispatcher prioritizations did have a statistically significant predictive value for critical care and two-day mortality, their discrimination was poor in comparison with all other risk assessment instruments with regards to hospital outcomes. This may in part be due to dispatchers prioritizing ambulance responses with an eye to the need for prehospital rather than in-hospital care. These aspects of patient care often coincide, but can in some cases differ. Cases of severe allergic reactions for instance call for a high priority ambulance response, but following treatment in the field by ambulance staff, often require only minimal in-hospital care. Effectively capturing this dimension of patient risk necessitates the definition of a different set of outcome measures than those reported here, and should be investigated in further studies.

We limited our analysis to hospital outcomes in order to allow for the direct comparison of models based on data collected at multiple points in the prehospital chain of care, and to facilitate comparison with other published research based on ED data. We also considered outcome measures based on ambulance data to be at greater risk for bias, as we suspect that the behavior of ambulance nurses may to some extent be influenced by the triage decisions of dispatch nurses. It should also be noted that the inclusion criteria used in this study were restrictive in that they excluded patients left at the scene of the incident, and patients transported to non-ED destinations. Upon implementation of these methods, care must be taken to ensure that the criteria used to include patients in a training dataset results in a population of patients similar to those upon whom the risk assessment tools will be applied.

Our models generally had lower levels of overall predictive value than found in previous studies investigating these outcomes based on data collected at the ED. This could in part be explained by population differences, given that the population of ambulance-transported patients investigated here constitutes a sum-population of the highest-acuity patients cared for at the ED [52–54]. The population in this study for instance had an average rate of in-hospital mortality of 4%, compared with the 0.5% rate found by Levin et al. [26], while our hospital admission rate was 52% as compared with the 30% found by Hong et al. [27], both of whom studied the full population of ED patients. It is also the case that the data available in records of prehospital care tend to be less detailed, lacking granular information regarding for instance the patient’s past medical history and laboratory test results. Such data have been found to provide substantial improvements to patient outcome predictions [27,29]. This study demonstrates that despite these barriers, prehospital data does have value in predicting hospital outcomes. We identified no studies of ED triage models which included prehospital data and as such, we suggest that one avenue for improving the performance of in-hospital triage models may be to include variables drawn from dispatch and ambulance records.

In conclusion, these results demonstrate that machine learning offers a viable approach to improving the accuracy of prehospital risk assessments, both in relation to existing rule-based triage algorithms, and current practice. Further research should investigate if the inclusion of additional unstructured data such as free-text notes and dispatch center call recordings could further improve the predictive value of the models reported here. Studies to investigate the attitudes of care providers with regards to risk assessments using ML may also prove fruitful; while ML methods can provide prehospital care providers with a more accurate risk score, the lack of direct interpretability often associated with such models may prove to be a barrier to acceptance. This study establishes only the feasibility of this approach to prehospital risk assessment, and further studies must establish the ability to influence the decisions of care providers and impact patient outcomes in prehospital care by means of prospective, preferably randomized, trial.

## Data Availability

Our ethics approval limits us to the publication of results at the aggregate level only, precluding us from publishing individual-level patient data. The Swedish Data Protection Authority has furthermore not yet endorsed a process for the anonymization of individually identifiable data which could be applied to ensure compliance with the EU General Data Protection Regulation in publishing this type of sensitive data. Data underlying the results are owned by the Uppsala Ambulance Service, and are available for researchers who meet the criteria for access to confidential data. Please contact the Uppsala Ambulance Service at to arrange access to the data underlying this study.

## Supporting Information

**S1 Table.** Predictor description. Descriptions of each set of predictors included in gradient boosting models, providing information regarding the number of non-missing, non-zero values among included calls, the average gain provided by the predictor, and the number of dummy-encoded variables included from the predictor in the models.

| Feature | Number of included calls with non-zero/non-missing value | Percent of included calls with non-zero/non-missing value | Average gain from inclusion of variables in ambulance models | Number of variables from group included in any ambulance model |
| --- | --- | --- | --- | --- |
| Ambulance - Airway findings | 34,719 | 90.9 | 0.212 | 4 |
| Ambulance - Pre-arrival notification given | 5,986 | 15.7 | 1.270 | 1 |
| Ambulance - Any intervention provided | 29,529 | 77.3 | 0.188 | 1 |
| Ambulance - Breathing findings | 34,599 | 90.6 | 0.325 | 8 |
| Ambulance - Breathing sounds | 31,482 | 82.4 | 0.132 | 3 |
| Ambulance - Call types | 31,239 | 81.8 | 0.748 | 26 |
| Ambulance - Circulation findings | 32,393 | 84.8 | 0.094 | 3 |
| Ambulance - CPR administered | 55 | 0.1 | 0.297 | 1 |
| Ambulance - Critical patient status | 3,267 | 8.6 | 0.377 | 1 |
| Ambulance - Time to dispatch | 19,815 | 51.9 | 1.630 | 1 |
| Ambulance - 12-lead EKG taken/sent to CICU | 9,128 | 23.9 | 0.335 | 1 |
| Ambulance - Patient immobilized | 764 | 2.0 | 0.112 | 1 |
| Ambulance - IV placed | 24,449 | 64.0 | 0.402 | 1 |
| Ambulance - Patient medical history | 16,130 | 42.2 | 0.315 | 7 |
| Ambulance - Medications administered | 22,365 | 58.5 | 1.208 | 14 |
| Ambulance - Oxygen administered (LPM) | 6,522 | 17.1 | 13.909 | 1 |
| Ambulance - Lights & siren to hospital | 5,140 | 13.5 | 4.307 | 1 |
| Ambulance - Priority to scene | 38,203 | 100.0 | 0.289 | 1 |
| Ambulance - Patient medications | 8,317 | 21.8 | 0.325 | 15 |
| Ambulance - Pulse quality | 33,019 | 86.4 | 0.577 | 6 |
| Ambulance - Time on scene | 35,315 | 92.4 | 2.887 | 1 |
| Ambulance - Skin condition | 29,858 | 78.2 | 0.121 | 3 |
| Ambulance - Time to hospital | 35,624 | 93.2 | 3.098 | 1 |
| Ambulance - AVPU | 37,172 | 97.3 | 1.212 | 1 |

| Feature | Number of<br>included calls<br>with non-zero/non-missing value | Percent of<br>included calls<br>with non-zero/non-missing value | Average gain<br>from inclusion<br>of variables in<br>ambulance<br>models | Number of<br>variables from<br>group included in<br>any ambulance<br>model |
| --- | --- | --- | --- | --- |
| Ambulance - Systolic blood pressure | 37,710 | 98.7 | 6.932 | 1 |
| Ambulance - Respiration rate | 36,567 | 95.7 | 4.824 | 1 |
| Ambulance - GCS | 36,985 | 96.8 | 6.292 | 1 |
| Ambulance - Pulse rate | 37,755 | 98.8 | 3.356 | 1 |
| Ambulance - SpO2 | 38,028 | 99.5 | 8.647 | 1 |
| Ambulance - Temperature | 32,473 | 85.0 | 5.047 | 1 |
| Dispatch - Patient Age | 38,203 | 100.0 | 12.050 | 1 |
| Dispatch - CDSS category | 38,203 | 100.0 | 4.300 | 32 |
| Dispatch - Distance to nearest ED | 38,141 | 99.8 | 3.256 | 1 |
| Dispatch - Patient Gender | 19,814 | 51.9 | 0.300 | 1 |
| Dispatch - Hour of call | 37,070 | 97.0 | 1.563 | 1 |
| Dispatch - Hours since last contact | 38,203 | 100.0 | 0.421 | 1 |
| Dispatch - Number of prior contacts (30 days) | 4,906 | 12.8 | 0.228 | 1 |
| Dispatch - Month of call | 38,203 | 100.0 | 0.901 | 1 |
| Dispatch - Number of CDSS questions answered | 37,374 | 97.8 | 1.409 | 1 |
| Dispatch - CDSS questions | 37,151 | 97.2 | 5.696 | 166 |
| Dispatch - CDSS recommended priority | 38,203 | 100.0 | 0.406 | 1 |

**S2 Analysis.**
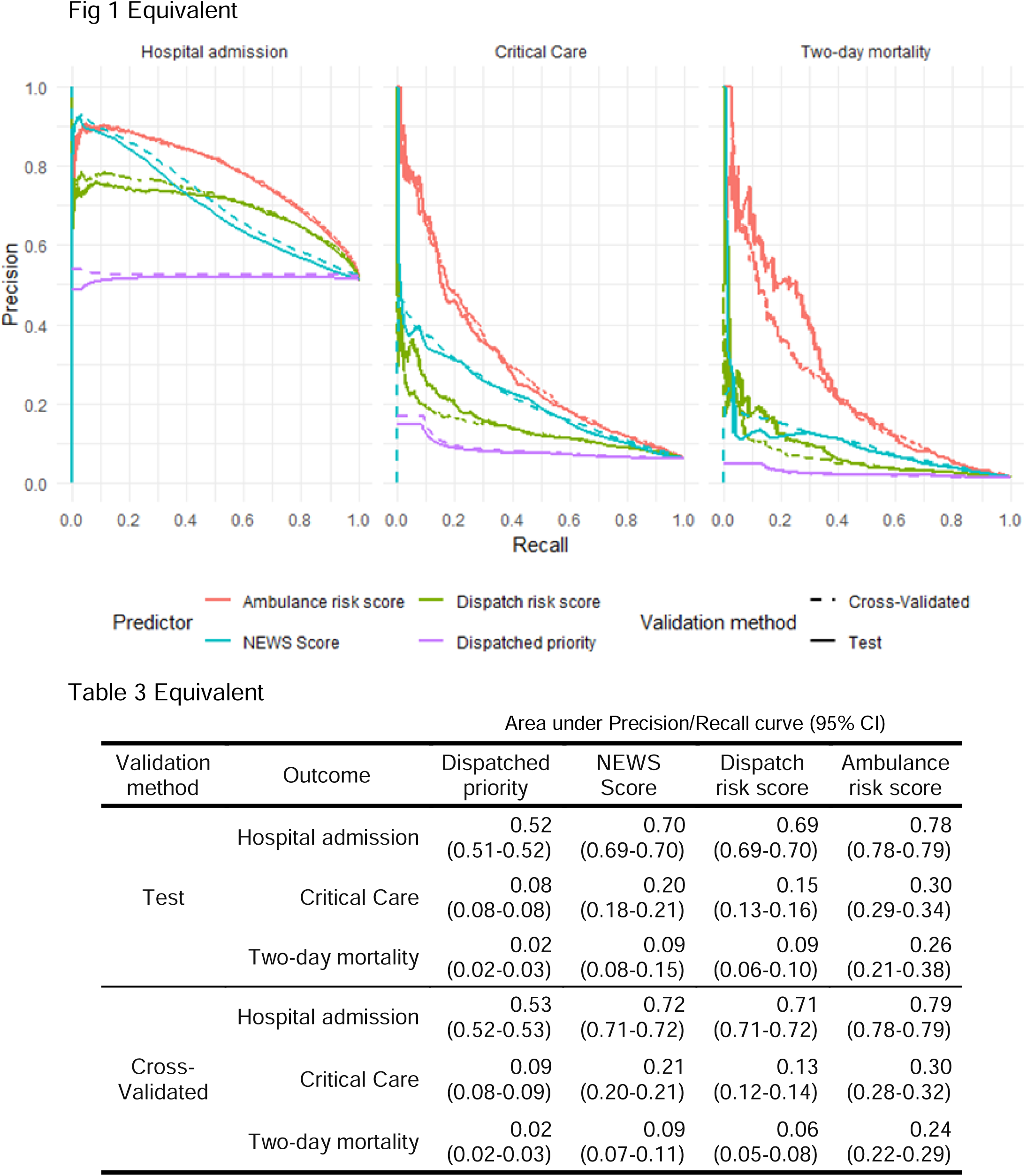
Precision/Recall analysis. Provides results from a Precision/recall curve analysis as commonly reported in the machine learning literature, presented in the same manner as Figure1 and table 2 in the main analysis.

**S3 Fig.**
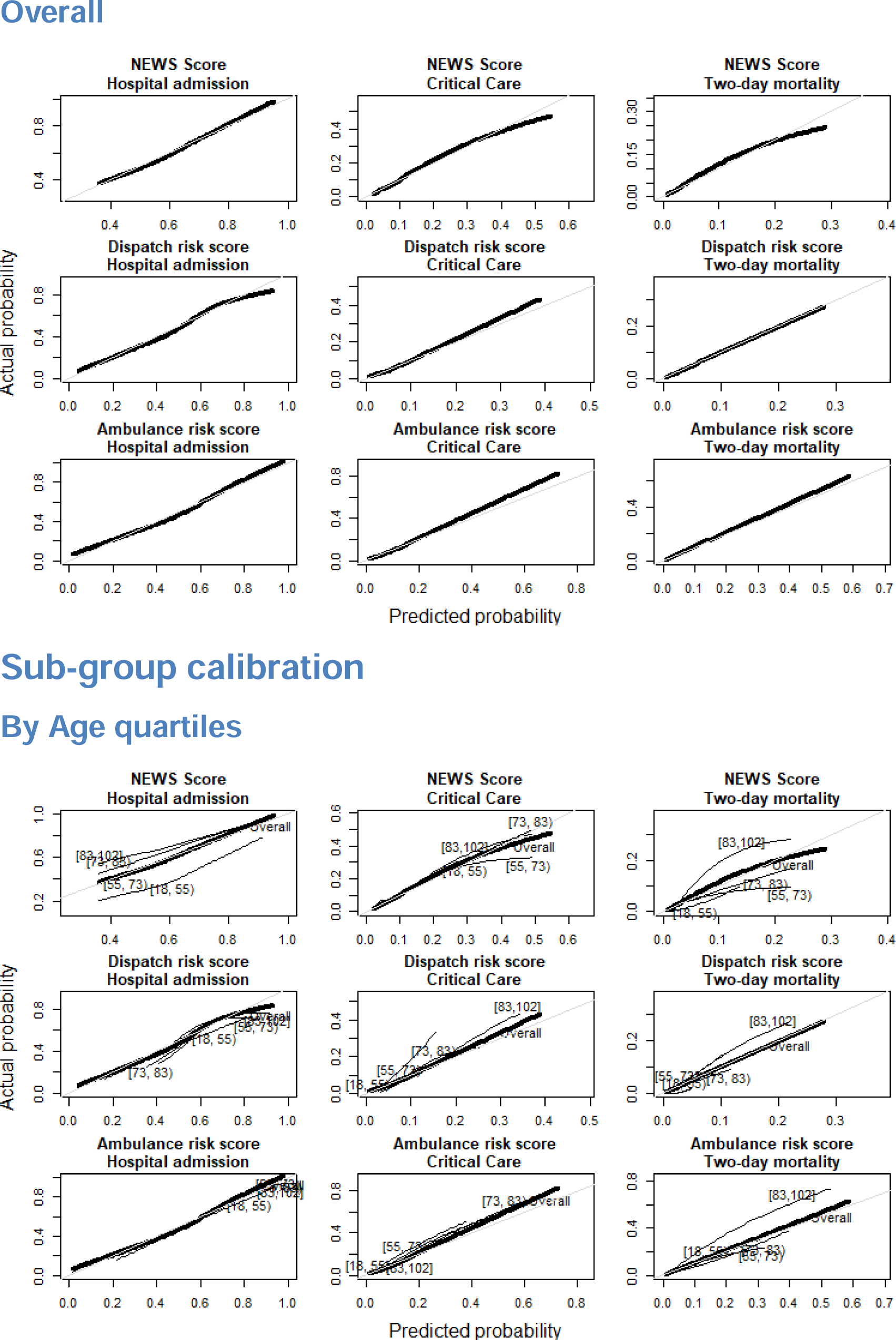
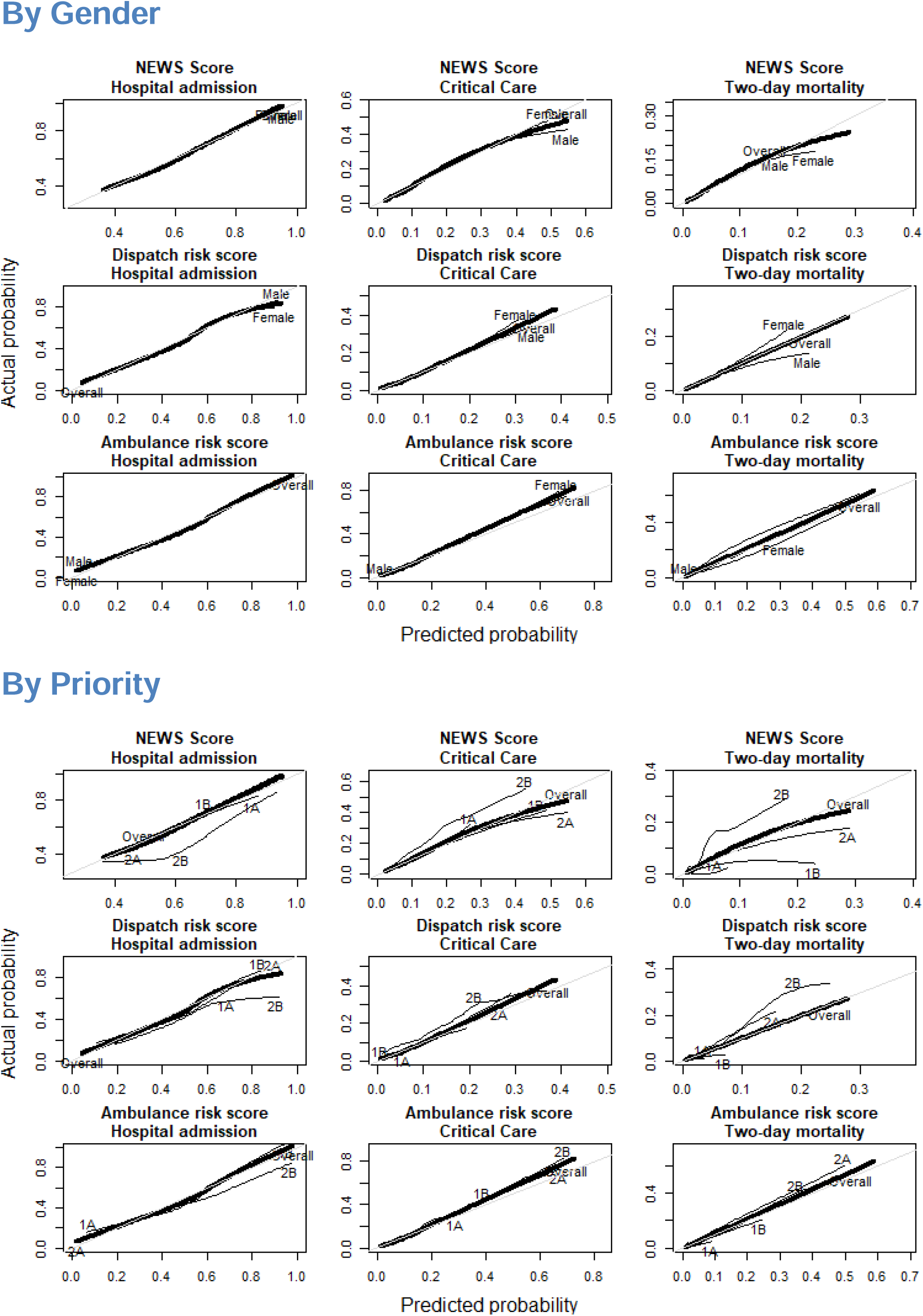
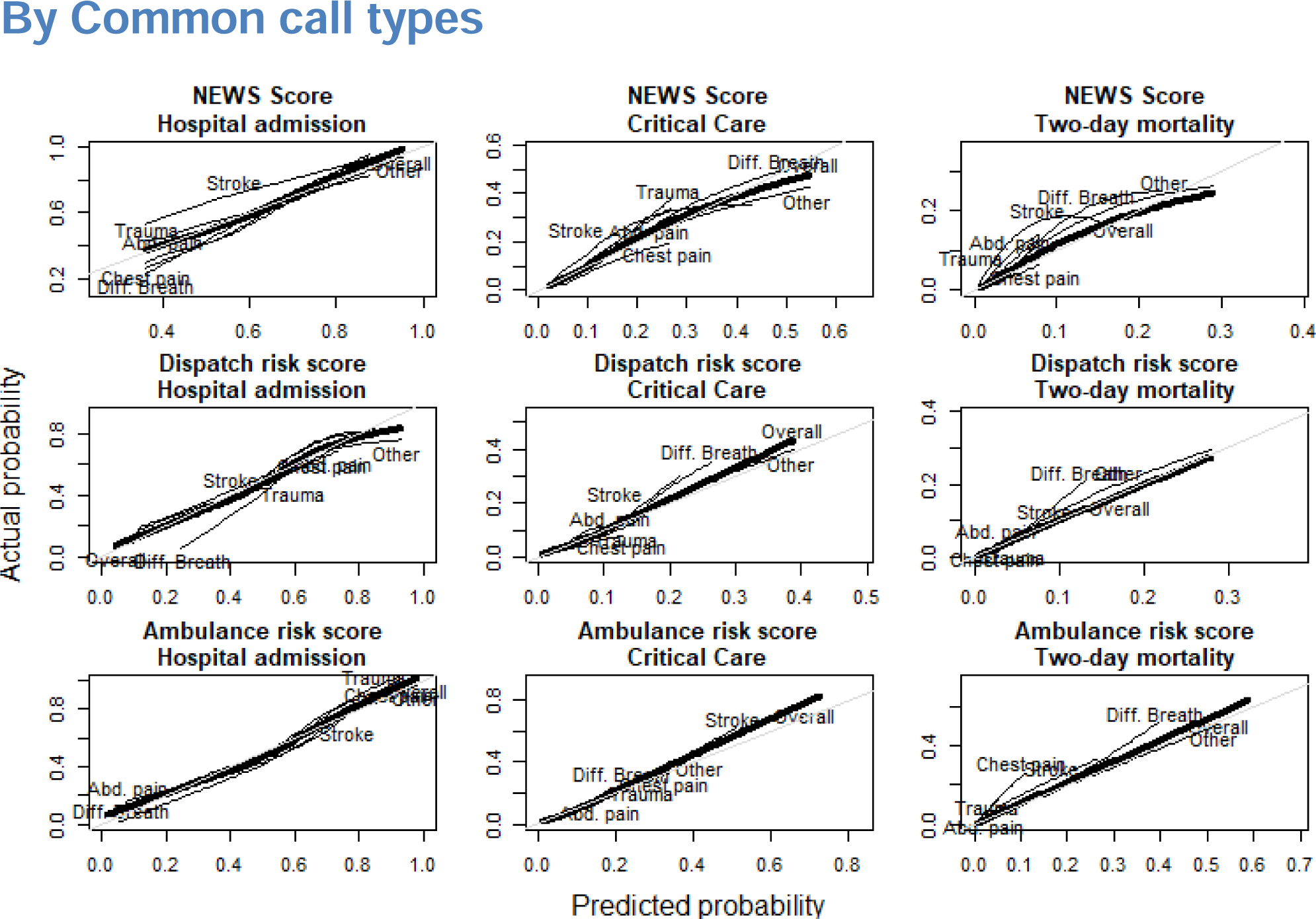
Model calibration curves. Provides the results of model calibration analyses using lowess smoothed calibration curves for both overall calibration, and calibration among sub-populations divided by age quartile, gender, call priority, and the 5 most common call types.

**S4 Table.** Model calibration mean average error. Provides summary statistics in the form of the mean average calibration error for NEWS and ML risk scores both in the full population, and the weighted average of all investigated sub-populations.

| Stratification variable | Predictor | Mean absolute error from ideal calibration |  |  |
| --- | --- | --- | --- | --- |
|  |  | Hospital admission | Critical Care | Two-day mortality |
| Overall | NEWS Score | 0.0112 | 0.0029 | 0.0016 |
|  | Dispatch risk score | 0.0170 | 0.0040 | 0.0009 |
|  | Ambulance risk score | 0.0228 | 0.0066 | 0.0011 |
| Age | NEWS Score | 0.0558 | 0.0064 | 0.0033 |
|  | Dispatch risk score | 0.0238 | 0.0079 | 0.0021 |
|  | Ambulance risk score | 0.0256 | 0.0093 | 0.0023 |
| Gender | NEWS Score | 0.0113 | 0.0055 | 0.0018 |
|  | Dispatch risk score | 0.0172 | 0.0055 | 0.0013 |
|  | Ambulance risk score | 0.0229 | 0.0077 | 0.0019 |
| Priority | NEWS Score | 0.0170 | 0.0045 | 0.0030 |
|  | Dispatch risk score | 0.0214 | 0.0061 | 0.0028 |
|  | Ambulance risk score | 0.0253 | 0.0071 | 0.0021 |
| Call type | NEWS Score | 0.0351 | 0.0068 | 0.0034 |
|  | Dispatch risk score | 0.0245 | 0.0067 | 0.0019 |
|  | Ambulance risk score | 0.0266 | 0.0080 | 0.0024 |

**S5 Table.** Sensitivity to alternate weights. Reports c-indexes for risk scores across a range of alternate weighting schemes, including the performance of individual model predictions across all investigated outcomes.

| Predictor set | Weights* | Hospital admission | Critical Care | Two-day mortality |
| --- | --- | --- | --- | --- |
| Dispatch | 100:10:1 | 0.74<br>(0.73-0.74) | 0.68<br>(0.67-0.69) | 0.75<br>(0.71-0.77) |
|  | 4:2:1 | 0.73<br>(0.73-0.74) | 0.70<br>(0.68-0.72) | 0.78<br>(0.75-0.81) |
|  | 1:1:1 | 0.72<br>(0.72-0.73) | 0.71<br>(0.70-0.72) | 0.79<br>(0.76-0.82) |
|  | 1:2:4 | 0.71<br>(0.71-0.72) | 0.70<br>(0.69-0.72) | 0.79<br>(0.77-0.82) |
|  | 1:10:100 | 0.70<br>(0.69-0.71) | 0.68<br>(0.67-0.69) | 0.79<br>(0.74-0.81) |
|  | 1:0:0 | 0.74<br>(0.73-0.75) | 0.66<br>(0.64-0.68) | 0.72<br>(0.70-0.74) |
|  | 0:1:0 | 0.68<br>(0.68-0.69) | 0.72<br>(0.70-0.74) | 0.78<br>(0.75-0.82) |
|  | 0:0:1 | 0.67<br>(0.67-0.68) | 0.65<br>(0.63-0.66) | 0.78<br>(0.75-0.80) |
| Ambulance | 100:10:1 | 0.79<br>(0.79-0.80) | 0.77<br>(0.76-0.78) | 0.86<br>(0.84-0.89) |
|  | 4:2:1 | 0.79<br>(0.78-0.80) | 0.78<br>(0.78-0.80) | 0.89<br>(0.87-0.91) |
|  | 1:1:1 | 0.79<br>(0.78-0.79) | 0.79<br>(0.78-0.80) | 0.89<br>(0.86-0.91) |
|  | 1:2:4 | 0.78<br>(0.77-0.78) | 0.80<br>(0.79-0.81) | 0.90<br>(0.88-0.92) |
|  | 1:10:100 | 0.78<br>(0.77-0.78) | 0.79<br>(0.77-0.81) | 0.89<br>(0.87-0.91) |
|  | 1:0:0 | 0.79<br>(0.79-0.80) | 0.75<br>(0.74-0.77) | 0.83<br>(0.80-0.86) |
|  | 0:1:0 | 0.73<br>(0.73-0.74) | 0.80<br>(0.80-0.82) | 0.90<br>(0.88-0.92) |
|  | 0:0:1 | 0.72<br>(0.71-0.72) | 0.76<br>(0.74-0.77) | 0.88<br>(0.87-0.90) |

\* Weights applied to model predictions for Hospital Admission : Critical Care : Two-day
Mortality

**S6 Code. R Source code**

Provides all R code necessary to replicate the results reported in this manuscript in a user-provided dataset. If no dataset is provided, results are calculated in a randomly generated synthetic dataset mimicking the univariate properties of our data. Be aware that the instruments will demonstrate essentially no predictive power if data is not provided. **Note to editor:** Upon publication, a link to a github repository containing a maintained version of the code will be placed here. **Note to preprint readers:** We’ll be releasing the source code upon publication – Who knows if some eagle-eyed reviewer will spot some error?

## References

1. Platts-Mills TF, Leacock B, Cabañas JG, Shofer FS, McLean SA. Emergency Medical Services Use by the Elderly: Analysis of a Statewide Database. Prehospital Emergency Care. 2010;14: 329–333. doi:10.3109/10903127.2010.481759

2. Lowthian JA, Jolley DJ, Curtis AJ, Currell A, Cameron PA, Stoelwinder JU, et al. The challenges of population ageing: Accelerating demand for emergency ambulance services by older patients, 1995–2015. Medical Journal of Australia. 2011;194. Available: https://www.mja.com.au/journal/2011/194/11/challenges-population-ageing-accelerating-demand-emergency-ambulance-services?inline=true

3. Hwang U, Shah MN, Han JH, Carpenter CR, Siu AL, Adams JG. Transforming Emergency Care For Older Adults. Health Affairs. 2013;32: 2116–2121. doi:10.1377/hlthaff.2013.0670

4. Pines JM, Mullins PM, Cooper JK, Feng LB, Roth KE. National Trends in Emergency Department Use, Care Patterns, and Quality of Care of Older Adults in the United States. Journal of the American Geriatrics Society. 2013;61: 12–17. doi:10.1111/jgs.12072

5. Dale J, Higgins J, Williams S, Foster T, Snooks H, Crouch R, et al. Computer assisted assessment and advice for “non-serious” 999 ambulance service callers: The potential impact on ambulance despatch. Emergency Medicine Journal. 2003;20: 178–183. doi:10.1136/emj.20.2.178

6. Haines CJ, Lutes RE, Blaser M, Christopher NC. Paramedic Initiated Non-Transport of Pediatric Patients. Prehospital Emergency Care. 2006;10: 213–219. doi:10.1080/10903120500541308

7. Gray JT, Wardrope J. Introduction of non-transport guidelines into an ambulance service: A retrospective review. Emergency Medicine Journal : EMJ. 2007;24: 727–729. doi:10.1136/emj.2007.048850

8. Magnusson C, Källenius C, Knutsson S, Herlitz J, Axelsson C. Pre-hospital assessment by a single responder: The Swedish ambulance nurse in a new role: A pilot study. International Emergency Nursing. 2015; doi:10.1016/j.ienj.2015.09.001

9. Krumperman K, Weiss S, Fullerton L. Two Types of Prehospital Systems Interventions that Triage Low-Acuity Patients to Alternative Sites of Care. Southern Medical Journal. 2015;108: 381–386. doi:10.14423/SMJ.0000000000000303

10. Eastwood K, Morgans A, Smith K, Hodgkinson A, Becker G, Stoelwinder J. A novel approach for managing the growing demand for ambulance services by low-acuity patients. Australian Health Review: A Publication of the Australian Hospital Association. 2015; doi:10.1071/AH15134

11. Höglund E, Schröder A, Möller M, Andersson-Hagiwara M, Ohlsson-Nevo E. The ambulance nurse experiences of non-conveying patients. Journal of Clinical Nursing. 2019;28: 235–244. doi:10.1111/jocn.14626

12. Kirkland SW, Soleimani A, Rowe BH, Newton AS. A systematic review examining the impact of redirecting low-acuity patients seeking emergency department care: Is the juice worth the squeeze? Emerg Med J. 2019;36: 97–106. doi:10.1136/emermed-2017-207045

13. Heward A, Damiani M, Hartley-Sharpe C. Does the use of the Advanced Medical Priority Dispatch System affect cardiac arrest detection? Emergency Medicine Journal. 2004;21: 115–118. doi:10.1136/emj.2003.006940

14. Bolorunduro OB, Villegas C, Oyetunji TA, Haut ER, Stevens KA, Chang DC, et al. Validating the Injury Severity Score (ISS) in different populations: ISS predicts mortality better among Hispanics and females. The Journal of Surgical Research. 2011;166: 40–44. doi:10.1016/j.jss.2010.04.012

15. Maddali A, Razack FA, Cattamanchi S, Ramakrishnan TV. Validation of the Cincinnati Prehospital Stroke Scale. Journal of Emergencies, Trauma, and Shock. 2018;11: 111–114. doi:10.4103/JETS.JETS_8_17

16. Silcock DJ, Corfield AR, Gowens PA, Rooney KD. Validation of the National Early Warning Score in the prehospital setting. Resuscitation. 2015;89: 31–35. doi:10.1016/j.resuscitation.2014.12.029

17. Seymour CW, Kahn JM, Cooke CR, Watkins TR, Heckbert SR, Rea TD. Prediction of Critical Illness During Out-of-Hospital Emergency Care. JAMA. 2010;304: 747–754. doi:10.1001/jama.2010.1140

18. Lane DJ, Wunsch H, Saskin R, Cheskes S, Lin S, Morrison LJ, et al. Assessing Severity of Illness in Patients Transported to Hospital by Paramedics: External Validation of 3 Prognostic Scores. Prehospital Emergency Care. 2019;0: 1–9. doi:10.1080/10903127.2019.1632998

19. Pirneskoski J, Kuisma M, Olkkola KT, Nurmi J. Prehospital National Early Warning Score predicts early mortality. Acta Anaesthesiologica Scandinavica. 2019;63: 676–683. doi:10.1111/aas.13310

20. Hettinger AZ, Cushman JT, Shah MN, Noyes K. Emergency Medical Dispatch Codes Association with Emergency Department Outcomes. Prehospital Emergency Care. 2013;17: 29–37. doi:10.3109/10903127.2012.710716

21. Veen M van, Steyerberg EW, Ruige M, Meurs AHJ van, Roukema J, Lei J van der, et al. Manchester triage system in paediatric emergency care: Prospective observational study. BMJ. 2008;337: a1501. doi:10.1136/bmj.a1501

22. Khorram-Manesh A, Montán KL, Hedelin A, Kihlgren M, Örtenwall P. Prehospital triage, discrepancy in priority-setting between emergency medical dispatch centre and ambulance crews.European Journal of Trauma and Emergency Surgery. 2010;37: 73–78. doi:10.1007/s00068-010-0022-0

23. Dami F, Golay C, Pasquier M, Fuchs V, Carron P-N, Hugli O. Prehospital triage accuracy in a criteria based dispatch centre. BMC Emergency Medicine. 2015;15. doi:10.1186/s12873-015-0058-x

24. Newgard CD, Yang Z, Nishijima D, McConnell KJ, Trent SA, Holmes JF, et al. Cost-Effectiveness of Field Trauma Triage among Injured Adults Served by Emergency Medical Services. Journal of the American College of Surgeons. 2016;222: 1125–1137. doi:10.1016/j.jamcollsurg.2016.02.014

25. Surgeons AC of. Resources for optimal care of the injured patient. 6th ed. chicago, IL; 2014.

26. Levin S, Toerper M, Hamrock E, Hinson JS, Barnes S, Gardner H, et al. Machine-Learning-Based Electronic Triage More Accurately Differentiates Patients With Respect to Clinical Outcomes Compared With the Emergency Severity Index. Annals of Emergency Medicine. 2018;71: 565–574.e2. doi:10.1016/j.annemergmed.2017.08.005

27. Hong WS, Haimovich AD, Taylor RA. Predicting hospital admission at emergency department triage using machine learning. PLOS ONE. 2018;13: e0201016. doi:10.1371/journal.pone.0201016

28. Raita Y, Goto T, Faridi MK, Brown DFM, Camargo CA, Hasegawa K. Emergency department triage prediction of clinical outcomes using machine learning models. Critical Care. 2019;23: 64. doi:10.1186/s13054-019-2351-7

29. Rajkomar A, Oren E, Chen K, Dai AM, Hajaj N, Hardt M, et al. Scalable and accurate deep learning with electronic health records. npj Digital Medicine. 2018;1: 18. doi:10.1038/s41746-018-0029-1

30. Blomberg SN, Folke F, Ersbøll AK, Christensen HC, Torp-Pedersen C, Sayre MR, et al. Machine learning as a supportive tool to recognize cardiac arrest in emergency calls. Resuscitation. 2019;138: 322–329. doi:10.1016/j.resuscitation.2019.01.015

31. Guttmann A, Schull MJ, Vermeulen MJ, Stukel TA. Association between waiting times and short term mortality and hospital admission after departure from emergency department: Population based cohort study from Ontario, Canada. BMJ. 2011;342: d2983. doi:10.1136/bmj.d2983

32. Di Somma S, Paladino L, Vaughan L, Lalle I, Magrini L, Magnanti M. Overcrowding in emergency department: An international issue. Internal and Emergency Medicine. 2015;10: 171–175. doi:10.1007/s11739-014-1154-8

33. Berg LM, Ehrenberg A, Florin J, Östergren J, Discacciati A, Göransson KE. Associations Between Crowding and Ten-Day Mortality Among Patients Allocated Lower Triage Acuity Levels Without Need of Acute Hospital Care on Departure From the Emergency Department. Annals of Emergency Medicine. 2019;74: 345–356. doi:10.1016/j.annemergmed.2019.04.012

34. Dugas AF, Kirsch TD, Toerper M, Korley F, Yenokyan G, France D, et al. An Electronic Emergency Triage System to Improve Patient Distribution by Critical Outcomes. The Journal of Emergency Medicine. 2016;50: 910–918. doi:10.1016/j.jemermed.2016.02.026

35. Newgard CD. The Validity of Using Multiple Imputation for Missing Out-of-hospital Data in a State Trauma Registry. Academic Emergency Medicine. 2006;13: 314–324. doi:10.1197/j.aem.2005.09.011

36. Laudermilch DJ, Schiff MA, Nathens AB, Rosengart MR. Lack of Emergency Medical Services Documentation Is Associated with Poor Patient Outcomes: A Validation of Audit Filters for Prehospital Trauma Care. Journal of the American College of Surgeons. 2010;210: 220–227. doi:10.1016/j.jamcollsurg.2009.10.008

37. Buuren S van, Groothuis-Oudshoorn K. Multivariate Imputation by Chained Equations in R. Journal of Statistical Software. 2011;45. Available: https://www.jstatsoft.org/article/view/v045i03

38. Chen T, Guestrin C. XGBoost: A Scalable Tree Boosting System. Proceedings of the 22Nd ACM SIGKDD International Conference on Knowledge Discovery and Data Mining. New York, NY, USA: ACM; 2016. pp. 785–794. doi:10.1145/2939672.2939785

39. Friedman J, Hastie T, Tibshirani R. The elements of statistical learning. Springer series in statistics New York; 2001.

40. Davison AC, Hinkley DV. Bootstrap Methods and Their Applications [Internet]. Cambridge: Cambridge University Press; 1997. Available: http://statwww.epfl.ch/davison/BMA/

41. Harrell FE. Rms: Regression Modeling Strategies [Internet]. 2017. Available: https://CRAN.R-project.org/package=rms

42. R Core Team. R: A Language and Environment for Statistical Computing [Internet]. Vienna, Austria: R Foundation for Statistical Computing; 2019. Available: https://www.R-project.org/

43. Cox S, Smith K, Currell A, Harriss L, Barger B, Cameron P. Differentiation of confirmed major trauma patients and potential major trauma patients using pre-hospital trauma triage criteria. Injury. 2011;42: 889–895. doi:10.1016/j.injury.2010.03.035

44. Fosbøl EL, Granger CB, Peterson ED, Lin L, Lytle BL, Shofer FS, et al. Prehospital system delay in ST-segment elevation myocardial infarction care: A novel linkage of emergency medicine services and inhospital registry data. American Heart Journal. 2013;165: 363–370. doi:10.1016/j.ahj.2012.11.003

45. Crilly JL, O’Dwyer JA, O’Dwyer MA, Lind JF, Peters JAL, Tippett VC, et al. Linking ambulance, emergency department and hospital admissions data: Understanding the emergency journey. Medical Journal of Australia. 2011;194: S34–S37. doi:10.5694/j.1326-5377.2011.tb02941.x

46. Birk HO, Henriksen LO. Prehospital Interventions: On-scene-Time and Ambulance-Technicians’ Experience. Prehospital and Disaster Medicine. 2002;17: 167–169. doi:10.1017/S1049023X00000406

47. Hale KE, Gavin C, O’Driscoll BR. Audit of oxygen use in emergency ambulances and in a hospital emergency department. Emergency Medicine Journal. 2008;25: 773–776. doi:10.1136/emj.2008.059287

48. Collins GS, Reitsma JB, Altman DG, Moons KGM. Transparent Reporting of a multivariable prediction model for Individual Prognosis or Diagnosis (TRIPOD): The TRIPOD statement. Annals of Internal Medicine. 2015;162: 55–63. doi:10.7326/M14-0697

49. Swaminathan S, Qirko K, Smith T, Corcoran E, Wysham NG, Bazaz G, et al. A machine learning approach to triaging patients with chronic obstructive pulmonary disease. PLOS ONE. 2017;12: e0188532. doi:10.1371/journal.pone.0188532

50. Goto T, Camargo CA, Faridi MK, Yun BJ, Hasegawa K. Machine learning approaches for predicting disposition of asthma and COPD exacerbations in the ED. The American Journal of Emergency Medicine. 2018;36: 1650–1654. doi:10.1016/j.ajem.2018.06.062

51. Hong WS, Haimovich AD, Taylor RA. Predicting 72-hour and 9-day return to the emergency department using machine learning. JAMIA Open. 2019; doi:10.1093/jamiaopen/ooz019

52. Marinovich A, Afilalo J, Afilalo M, Colacone A, Unger B, Giguère C, et al. Impact of Ambulance Transportation on Resource Use in the Emergency Department. Academic Emergency Medicine. 2004;11: 312–315. doi:10.1111/j.1553-2712.2004.tb02218.x

53. Ruger JP, Richter CJ, Lewis LM. Clinical and Economic Factors Associated with Ambulance Use to the Emergency Department. Academic Emergency Medicine. 2006;13: 879–885. doi:10.1197/j.aem.2006.04.006

54. Squire BT, Tamayo A, Tamayo-Sarver JH. At-Risk Populations and the Critically Ill Rely Disproportionately on Ambulance Transport to Emergency Departments. Annals of Emergency Medicine. 2010;56: 341–347. doi:10.1016/j.annemergmed.2010.04.014

